# Vaccines, social measures and Covid19 - A European evidence-based analysis Vaccines, social measures and Covid19

**DOI:** 10.1101/2021.04.15.21255558

**Authors:** John R Porter

**Author notes:** Strandgaardsvej 32, 4000 Roskilde, Denmark.

## Abstract

**Background:** A fully quantitative picture of national effectiveness in controlling the spread of the Covid19 virus should consider the percentage of a population vaccinated in relation to the percentage of a population as active cases.

**Methods:** Publicly available data from 27 European countries on nine dates in 2021. Data were (i) initial Covid19 vaccinations and (ii) Covid19 active cases, both as percentages of a country’s population. Dividing (i) by (ii) yielded a new metric, the V ratio, which can increase as (i) increase or as (ii) decreases or both. I correlated the change in V ratio with the change in R statistic in the 27 counties and nine dates.

**Results:** Mean European V ratio increased from January 11 2021 onwards; inverse correlation was found between V ratio and R statistic (p<0.001, r2=0.15, df=234). Initial threshold V ratio of 10-15 resulted in an R statistic of 1.0 or lower; this threshold increased to 30-40 with further vaccinations. Variation between countries in the V ratio increased with time.

**Conclusion:** This quantitative assessment and use of a summary data-derived threshold index showed the integrated effectiveness of vaccinations and social measures for European countries for Covid19. It established a threshold range for an R value of 1 and calculation of the number of vaccinations needed in Europe to reduce the infectivity of the virus to unity. Results can be used to quantify the relation between transmission following vaccination and social measures to control the spread of Covid19.

**Summary box:** ‘What is already known’ in the epidemiology of the Covid19 pandemic in Europe countries is time- and country-based estimates of the R statistic as a measure of infectivity, the percentages of vaccinated persons to reduce infectivity and the number of active cases amenable to social measures. What is not known is how these three parameters interact.

What does this study add’? This study adds by invention an index (V) of percentage vaccinated population divided by percentage active cases. It then examines the corresponding V index in relation to the R statistic. It derives a range threshold V ratio for an R statistic of unity and extends this to suggest the total number of vaccines needed in Europe.

**Policy implications:** These results will allow policy judgements to be made on the basis of measured evidence, and not just models, of the means to reduce the Covid19 pandemic in Europe. The R statistic captures the development of the pandemic; the V ratio measures the integrated and quantified measures to reduce it. Further development of the analysis could assist in calculating the relative effectiveness of vaccination and social measures linked to a range of values of the V ratio. It can also be used as an alarm call to identify states which, even given 100% vaccination, may not reduce their R statistic below unity.

## INTRODUCTION

Looking just at number of administered vaccines misses at least half the story of the European societal response to Covid19. To get a balanced picture, one has to consider the percentage of a country’s population vaccinated in relation to the percentage of a population as active cases. Reducing active cases via social measures is just as, and maybe more, effective as vaccinating. The history of this paper was an article in the Danish newspaper *Politiken*, in which they presented the percentage of populations in a range of European countries that had received their first Covid19 vaccination. My initial reaction was – that is all well and good but it is only telling half the story; the other half is what percentage of a national population can still be classed as being active cases. There is much discussion about the ‘race’ between the numbers of people being vaccinated against Covid19 in Europe and the need to reduce the number of active cases, which is highly responsive to social measures of restricting, or not, the disease’s, spread. The ‘race’ to reduce the number of people ill from the Covid19 pandemic has two main runners – the percentage of a population vaccinated and the percentage of a population as active cases, defined as the total of Covid19 clinical, laboratory and epidemiological data (https://www.ecdc.europa.eu/en/covid-19/surveillance/case-definition). Data on active cases come from https://www.worldometers.info/coronavirus/; data on vaccination rates come from https://ourworldindata.org/covid-vaccinations.

## MATERIALS AND METHODS

The R statistic is used to characterise the infection rate of the virus; a similar index could indicate the link between vaccinations and active cases at a point in time. I have called this index the ‘V ratio’ and it is formally defined *as the ratio of the* p*ercentage of a population first vaccinated divided by the percentage of a population infected as active cases*. If the ratio is larger than 1.0 then vaccinations are relatively running ahead of active cases and *vice versa*. The ratio will change over time and, as an example, on January 26 2021 (Table 1), the V ratio had a value in Denmark of 3.6/0.16 *ie* 23.14 and in the UK of 10.4/3.13, that is 3.32.

**Table 1.**
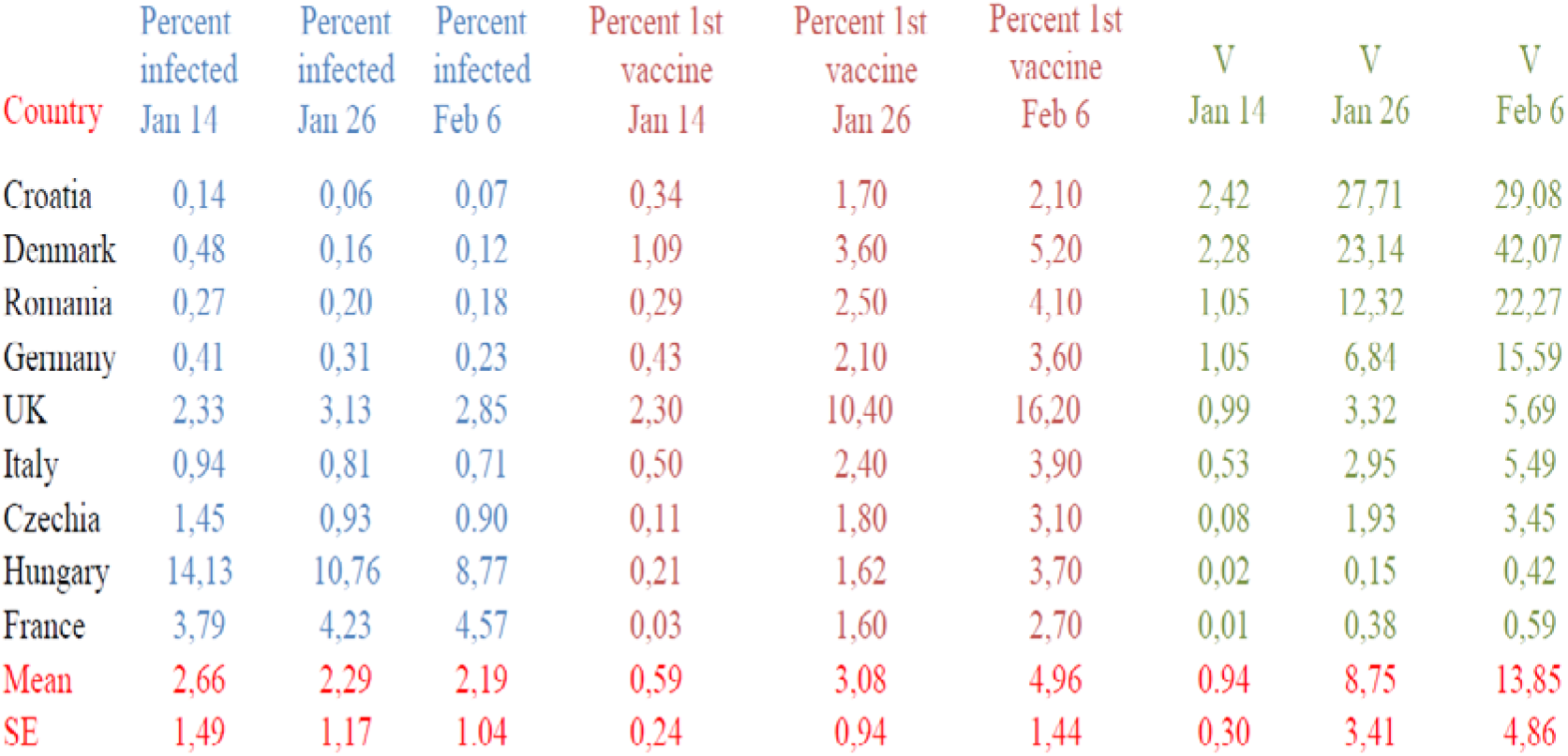
Percentage of population infected, received first vaccine and calculated V number (V) for a selection of EU member states and the UK for early stages of the vaccination programme with means and standard errors (SE). Data for all reporting EU and EEA member states are available from the author. Data sources - https://www.worldometers.info/coronavirus; https://ourworldindata.org/covid-vaccinations.

| Country | Percent infected<br>Jan 14 | Percent infected<br>Jan 26 | Percent infected<br>Feb 6 | Percent 1st vaccine<br>Jan 14 | Percent 1st vaccine<br>Jan 26 | Percent 1st vaccine<br>Feb 6 | V<br>Jan 14 | V<br>Jan 26 | V<br>Feb 6 |
| --- | --- | --- | --- | --- | --- | --- | --- | --- | --- |
| Croatia | 0,14 | 0,06 | 0,07 | 0,34 | 1,70 | 2,10 | 2,42 | 27,71 | 29,08 |
| Denmark | 0,48 | 0,16 | 0,12 | 1,09 | 3,60 | 5,20 | 2,28 | 23,14 | 42,07 |
| Romania | 0,27 | 0,20 | 0,18 | 0,29 | 2,50 | 4,10 | 1,05 | 12,32 | 22,27 |
| Germany | 0,41 | 0,31 | 0,23 | 0,43 | 2,10 | 3,60 | 1,05 | 6,84 | 15,59 |
| UK | 2,33 | 3,13 | 2,85 | 2,30 | 10,40 | 16,20 | 0,99 | 3,32 | 5,69 |
| Italy | 0,94 | 0,81 | 0,71 | 0,50 | 2,40 | 3,90 | 0,53 | 2,95 | 5,49 |
| Czechia | 1,45 | 0,93 | 0,90 | 0,11 | 1,80 | 3,10 | 0,08 | 1,93 | 3,45 |
| Hungary | 14,13 | 10,76 | 8,77 | 0,21 | 1,62 | 3,70 | 0,02 | 0,15 | 0,42 |
| France | 3,79 | 4,23 | 4,57 | 0,03 | 1,60 | 2,70 | 0,01 | 0,38 | 0,59 |
| Mean | 2,66 | 2,29 | 2,19 | 0,59 | 3,08 | 4,96 | 0,94 | 8,75 | 13,85 |
| SE | 1,49 | 1,17 | 1,04 | 0,24 | 0,94 | 1,44 | 0,30 | 3,41 | 4,86 |

By February 6 2021, their V ratios had increased to 42.07 and 5.69 respectively. Such analysis shows, for example, that the UK is percentage-wise more successful than Denmark as regards vaccination percentage, but Denmark had a higher V ratio because its active case percentage was more than 20 times lower than that in the UK. The V ratio integrates the relationship between vaccinations and active cases and, I submit, is a simple but useful summary statistic.

There are many ways to calculate the R statistic for infectivity and it has attracted criticism^1^ as not being the tool needed to manage the pandemic (https://www.nature.com/articles/d41586-020-02009); I calculated R simply as *ln* (active cases)/the interval between samples in days; thus it is an instantaneous value, but close to other reported values that take account of factors such as a country’s population age-distribution. The R statistic changes slowly over time – (www.gov.uk/guidance/the-r-number-in-the-uk) and it is important to remember, in the current context, that it is a measure of person-to-person infectivity and not a measure of the intrinsic rate of increase of the virus population, as would be defined in the original Lotka equation. The R statistic is mostly given as a range of value (https://plus.maths.org/content/epidemic-growth-rate). My calculated average R statistic of the European countries in the period from January 14 2021 to February 26 2021 were between 1.08 (sd. 0.20) and 0.88 (sd. 0.16), which are reasonable values. My calculation of the European average R statistic on March 10 2021 was 0.9 with a sd. of 0.15; also a reasonable value. Out of 26 country calculations I made of the R statistic, 22 were within the Poisson method R distributions given independently by http://metrics.covid19-analysis.org.

## RESULTS

On February 6 2021 for the presented EU countries and the UK (Table 1), the mean percentage active cases was 2.19%, the percentage vaccinated was 4.96%, giving an average European V ratio of 13.85, which was a creditable initial post-vaccine start for the EU, however the standard errors of these mean values are quite high, indicating considerable variation between European countries. By February 26 the average European V ratio had increased to 19.39, but with a standard difference of 20.84, indicating large intra-European variation, but without a clear geographical signal. Also, I have not accounted for the relative efficacy of vaccination in these calculations. This could be done by dividing the V ratio by 0.85 to account for an 85% effectiveness of the vaccines. Supplementary Material (V_number_jrp_210301.pdf) shows an example calculation file from February 26 2021.

Further work (Figure 1) shows a strong negative relation between R statistic which declines as the V ratio increases. This may allow an estimate to be made of when the R statistic will reduce to a safe level, following vaccination in a country or a region in a country. The R statistic value changes relatively slowly (www.gov.uk) but small changes have a large effect on total numbers of infections; the V ratio changes more quickly and can also have a large effect. Figure 1 shows for data from February 6 2021, as expected, that as the V ratio increases that the R statistic declines, but the most important point is the V ratio for an R statistic of 1.0, indicated on Figure 1 by the vertical red line. This line crosses the diagonal regression line at a V ratio (on the vertical axis) of about 10. The conclusion using data to that point was that if the percentage of a population that is vaccinated is more than about 10 times higher than the percentage of active cases, then there are grounds to expect that the R statistic will not go above 1.0. Taking a conservative account of a relative effectiveness of vaccines of 85% – a V ratio in excess of 12 would have been more advisable. As with the R statistic, the V ratio has a range of values. Supplementary Material (deltaR_deltaV_jrp_210227.pdf) in which I correlated the relationship between R and V ratio for consecutive time periods between dates in January and February 2021, termed delta-V and delta-R. This shows the delta-R value falling with date and increasing delta-V. Such interval analysis is a stringent test of the relationship between falling R value and rising V ratio. If the two methods of an interval analysis and single date analyses are leading to the same finding then this adds confidence to the robustness of conclusions. As a climate scientist who has been an author with the Intergovernmental Panel on Climate Change since 1995, I learned to appreciate the importance of multiple evidences^2^.

**Figure 1.**
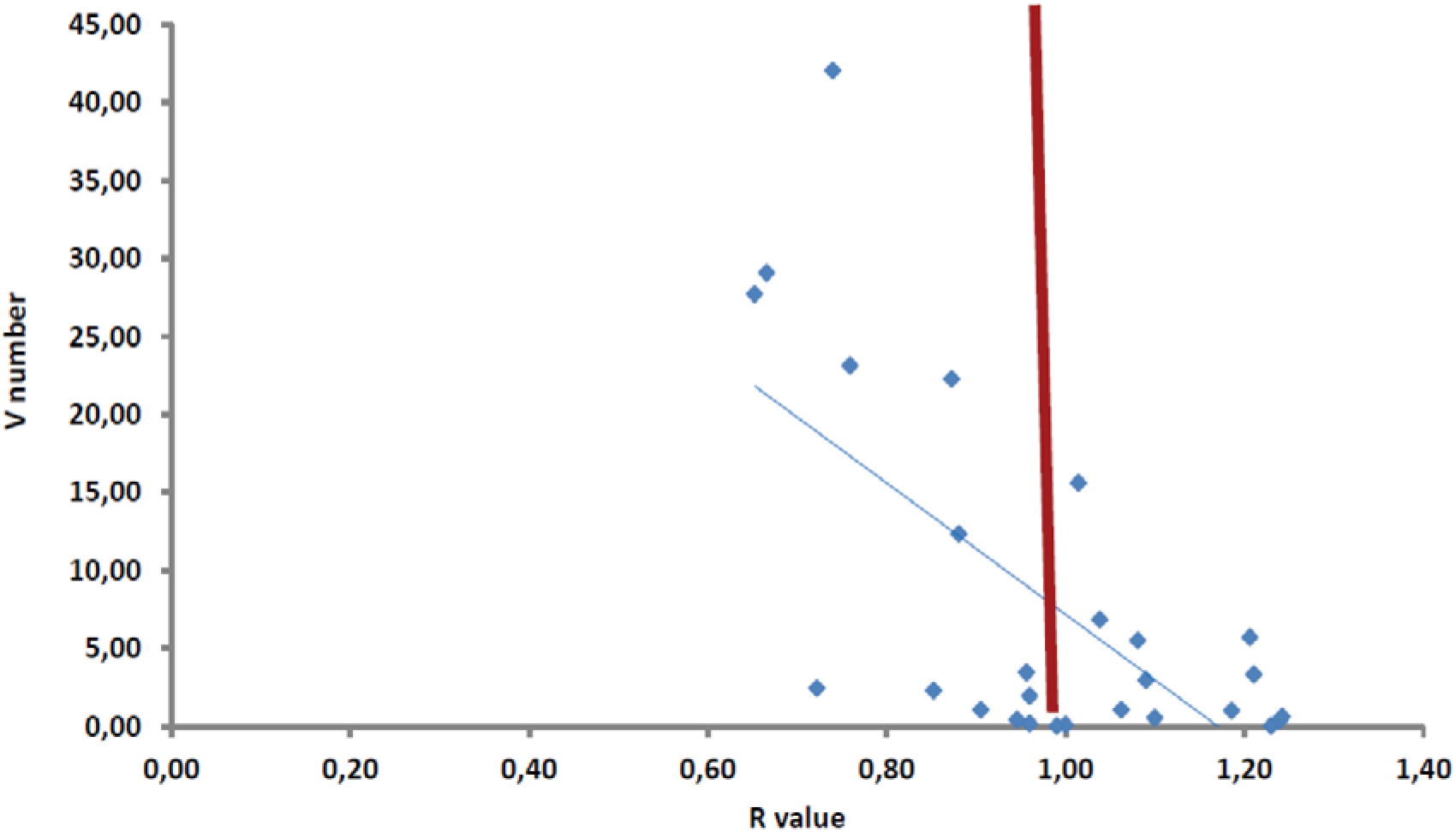
Relationship between the R statistic and the V ratio for the selected countries on February 6 2021 from Table 1. The vertical red line marks the R statistic with a value of 1.0.

However, February 6 was a relatively early date in the roll-out of vaccinations in European countries. A more robust assessment of the relation between V ratio and R statistic is seen from later data (Figure 2). These data are for 24 EU member states plus Norway, Switzerland and the UK and show a significant polynomial correlation between increasing V ratio and declining R statistic (p<0.001), although the coefficient of determination of the data is low, but that is of lesser importance for the study. The polynomial line of the quadratic relationship is in fact a timeline from low V ratio and high R statistic (bottom right) to high V ratio and lower R statistic (upper left), as the percentage of people vaccinated increases and the percentage of active cases, via social measures, falls. The increased spread in V ratio with time infers that countries have used different combinations of percentage vaccination and social measures to try and reduce the R statistic.

**Figure 2.**
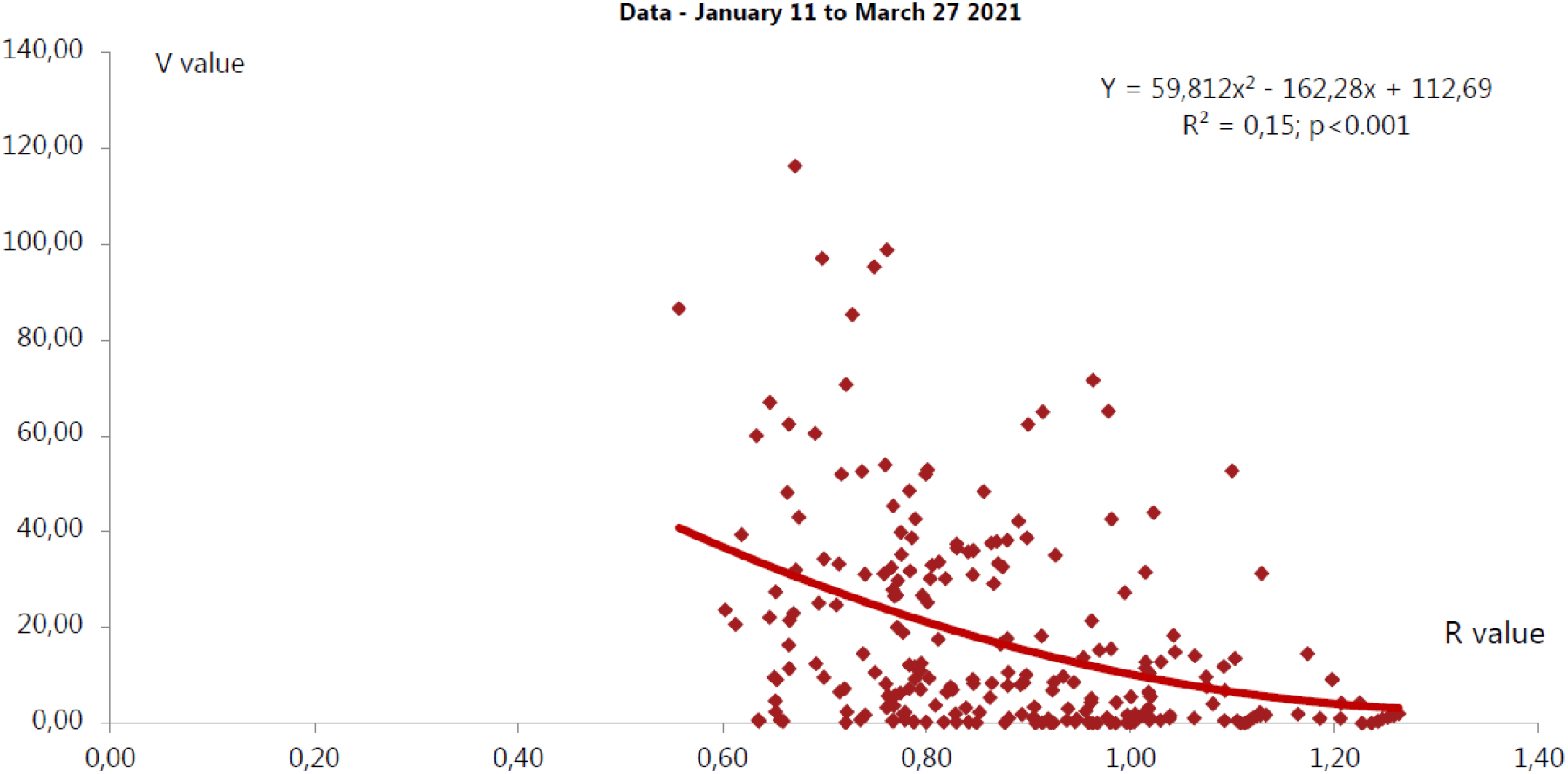
Relationship between the R statistic and the V ratio for 24 EU member states plus Norway, Switzerland and the UK from January 11 to March 27 2011.

## DISCUSSION

The V ratio emphasises the value there has been of preventative lock-down measures to limit virus spread. Until now, social prevention of disease spread has naturally played a larger role than vaccination but with a high and optimistic expectation that this balance will tip in favour of vaccination. R captures the increase and decrease of the pandemic’s spread; V captures the means, both social and medical, to control it. R portrays preventable spread, V defines a cure and with both factors operating, there is reason to be optimistic about limiting the disease to easily manageable levels, and in dealing with the pandemic, one has to include both preventative measures and curative ones. The V ratio presents both these aspects as a quantitative and simple index.

There are caveats of course – the R statistic could be calculated in a more nuanced way; the number of sample countries and frequency of sampling could be increased; the study could be extended outside Europe – but I do wish to make the point that all my calculations are based on empirical and publicly available data from official sources. The only modelling involved is my calculation of R – but my calculated numbers are conservative and close to observed, for example to the UK figures - www.gov.uk/guidance/the-r-number-in-the-uk.

A second caveat is that there could be an overlap between people who are active cases and people who are vaccinated – i.e. one could double count them. But is it both unlikely and also would not have much effect on the V ratio as the difference between the vaccination and active case percentages are large. So, if the active cases are 0.5% and the vaccination percentage is 5 then the V ratio is 10. If all the active cases are also in the vaccination percentage then the V ratio becomes 4.5%/0.5%, a value of nine, with little change in the V ratio. However, more nuanced calculation of the V ratio needs to take account of the demographics of different countries, whether active cases are a percentage of the total whole population or just of the population of potential active cases. However, such distinctions should not make major change to my essential message that there exists a range of V ratio values that inversely correlate with the R statistic and can be used to define useful parameters from readily available pandemic data.

Two other important issues can be investigated using the index and relationships presented in this paper; I hope others will take up the challenge. The first question is whether it has been vaccines or the social measures that have had the higher impact on any decline in active cases. One way to approach this would be to examine the changes in infections per day in a period prior to the commencement of vaccinations, perhaps in November 2020, and compare this with the rate of decline in infections per day in a period with both social measures and vaccinations, perhaps February 2021, - the difference between the two rates would then quantify the relative effectiveness of social preventative measures and the vaccine based attempt to cure the pandemic.

The second important question is whether vaccination reduces the infectivity of the virus – are people who have been vaccinated less likely to transmit the virus? The ideal data to answer such a question would be to find a period of time after vaccination started, and during which the vaccination percentage does not change, and then look at changes in the number of active cases – but for a period perhaps three weeks after the vaccination period as infection does not follow immediately from contact with the virus. The hypothesis would be that if previous vaccination reduces transmission then percentage of active cases following the vaccination period plus delay should fall.

To get an estimate of how many vaccinations are needed in the EU and also the degree to which vaccination on its own can reduce the R statistic to unity, I calculated, on the basis of a country’s V ratio in relation to one of 40, and scaled this to the proportion of the population vaccinated – so that should give the number of shots (corrected for double shots) for the V ratio to rise to 40. For example, country A could have a V ratio of 10 and have vaccinated 10% of its population. It has to quadruple its V ratio to approach an R of unity, but this is not a problem as it has 90% of the population un-vaccinated.

This calculation does not account for a decline in active cases via social distancing and isolation - so it represents the worst case scenario for the needed vaccination numbers. I calculate the worst case, as of March 21 2021, being 612 million vaccinations with the highest number for France. The reasoning for this calculation is as follows: the observed active cases percentage in Europe is just below 2%; to achieve a V ratio of 40 (Figure 2) thus requires a percentage vaccination of 80% and this is doubled for two vaccinations; should the percentage infection increase then the percentage vaccinations would also need to increase to preserve the V ratio at its threshold range of value. Vaccination rate has to be scaled from % per country to % per person by taking account of the total population, with the caveat that not all persons in a population (eg. children) are to be vaccinated and also that vaccinations are not 100% effective. This latter point carries an important message: if for example country A had a V ratio of 2, aiming for 40 – *i*.*e*. at 5% of the goal – but if, say, 10% of its population has been vaccinated, then the maximum possible V ratio in country A would be 20; thus the country would have to make as large an effort in isolation and social distancing as it has with vaccination.

## CONCLUSION

Many statements of the conditions for human societies to come out of restrictions imposed to limit the pandemic and the need to vaccinate are qualitative or semi-quantitative (i.e. more or less). I have tried to make quantitative and evidence based statements of the joint and relative effectiveness of social measures and vaccination, using publicly available data. I would strongly suggest that others check this work for other countries to see if there is a robust relation between the V ratio and R statistic. This is important quantitative information for policy people to help them in their decisions on the quantitative data-based conditions needed to take a country out of restrictions from the Covid19 pandemic. The evidence needs to be published in the scientific press and also publicly. (3030 words).

## Data Availability

Data are available on request to the author.

## Supplementary Materials

Figure

S1 - deltaR_deltaV_jrp_210227.pdf

Data File

S1 – V_ratio_jrp_210301.pdf

