## Supplement Table 1 for "Vaccines, social measures and Covid19 - A European evidence-based analysis Vaccines, social measures and Covid19"

| Countries | Active Cases | Population | Percent infected | Percent 1st vaccine | **V number (E/D)** | R value |
|---|---|---|---|---|---|---|
| Austria | 24090 | 9033299 | 0,27 | 9,55 | 35,81 | 0,84 |
| Belgium | 715494 | 11616051 | 6,16 | 8,55 | 1,39 | 1,12 |
| Bulgaria | 38385 | 6920829 | 0,55 | 4,35 | 7,84 | 0,88 |
| Croatia | 3981 | 4091912 | 0,10 | 5,89 | 60,54 | 0,69 |
| Czechia | 163812 | 10719408 | 1,53 | 8,43 | 5,52 | 1,00 |
| Danmark | 7977 | 5802895 | 0,14 | 13,11 | 95,37 | 0,75 |
| Estonia | 13764 | 1327004 | 1,04 | 11,31 | 10,90 | 0,79 |
| Finland | 17113 | 5545238 | 0,31 | 10,39 | 33,67 | 0,81 |
| France | 3604581 | 65349297 | 5,52 | 8,80 | 1,60 | 1,26 |
| Germany | 125432 | 83924420 | 0,15 | 9,74 | 65,17 | 0,98 |
| Greece | 25533 | 10396218 | 0,25 | 7,62 | 31,03 | 0,85 |
| Hungary | 123691 | 967440 | 12,79 | 14,76 | 1,15 | 0,98 |
| Ireland | 196725 | 4966548 | 3,96 | 10,59 | 2,67 | 1,02 |
| Italy | 487074 | 60415065 | 0,81 | 9,56 | 11,86 | 1,09 |
| Latvia | 8052 | 1875165 | 0,43 | 4,57 | 10,64 | 0,75 |
| Lithuania | 10556 | 2702157 | 0,39 | 11,64 | 29,80 | 0,77 |
| Luxembourg | 2930 | 631269 | 0,46 | 7,59 | 16,35 | 0,67 |
| Malta | 3182 | 442160 | 0,72 | 23,05 | 32,03 | 0,67 |
| Norway | 10277 | 545365 | 1,88 | 11,18 | 5,93 | 0,77 |
| Poland | 274411 | 37824814 | 0,73 | 10,80 | 14,89 | 1,04 |
| Portugal | 57152 | 10181057 | 0,56 | 10,24 | 18,24 | 0,91 |
| Romania | 48106 | 19169972 | 0,25 | 9,72 | 38,73 | 0,90 |
| Slovakia | 66147 | 5461031 | 1,21 | 10,42 | 8,60 | 0,92 |
| Slovenia | 10464 | 2079088 | 0,50 | 10,09 | 20,05 | 0,77 |
| Spain | 269823 | 47766667 | 0,56 | 10,37 | 18,36 | 1,04 |
| Switzerland | 32660 | 8690652 | 0,38 | 10,96 | 29,16 | 0,87 |
| UK | 761448 | 68099968 | 1,12 | 35,02 | 31,32 | 1,13 |
| mean | 263069 | 18020185 | 1,58 | 11,05 | 23,65 | 0,90 |
| sd | 699253 | 24600373 | 2,73 | 5,90 | 22,08 | 0,15 |
